# Ceramides are decreased after liraglutide treatment in two randomized clinical trials

**DOI:** 10.1101/2023.03.21.23287536

**Authors:** Asger Wretlind, Viktor Rotbain Curovic, Andressa de Zawadzki, Tommi Suvitaival, Jin Xu, Emilie Zobel, Bernt Johan von Scholten, Rasmus Sejersten Ripa, Andreas Kjaer, Tine Willum Hansen, Tina Vilsbøll, Henrik Vestergaard, Peter Rossing, Cristina Legido-Quigley

## Abstract

**Background:** Specific ceramides have been identified as risk markers for cardiovascular disease (CVD) years before onset of disease. Treatment with the glucagon-like peptide-1 receptor agonist (GLP-1RA) liraglutide has been shown to induce beneficial changes in the lipid profile and reduce the risk of CVD. Reducing lipotoxic lipids with an antidiabetic drug therapy could be a path towards precision medicine approaches for the treatment of complications to diabetes. In this *post-hoc* study, we investigated the effect of liraglutide on CVD-risk associated ceramides in two randomized clinical trials including participants with type 2 diabetes (T2D).

**Methods:** This study analyzed plasma samples from two independent randomized placebo-controlled clinical trials. The first trial, Antiproteinuric Effects of Liraglutide Treatment (LirAlbu12) followed a crossover design where 27 participants were treated for 12 weeks with either liraglutide (1.8 mg/d) or placebo, followed by a four-week washout period, and then another 12 weeks of the other treatment. The second clinical trial, Effect of Liraglutide on Vascular Inflammation in Type-2 Diabetes (LiraFlame26), lasted for 26 weeks and followed a parallel design, where 102 participants were randomized 1:1 to either liraglutide or placebo. Here we measured six prespecified plasma ceramides using liquid chromatography mass spectrometry and assessed their changes using linear mixed models and t-tests. Possible confounders were assessed with mediation analyses.

**Results:** In the LirAlbu12 trial, C16 Cer and C24 Cer were reduced with liraglutide treatment (p <0.05) compared to placebo. In the LiraFlame26 trial, treatment with liraglutide resulted in a significant reduction of two ceramides associated with CVD risk, C16 Cer and C24:1 Cer (p <0.05) compared to placebo. None of the remaining ceramides showed statistically significant changes in response to liraglutide treatment compared to placebo. Mediation analyses showed that weight loss did not affect ceramide reduction.

**Conclusions:** We demonstrated that treatment with liraglutide resulted in a consistent reduction in C16 Cer after both 12 and 26 weeks of treatment and an overall decreasing trend was observed for two other ceramides. Our findings suggest the GLP-1RA can be used also to modulate ceramides.

**Trial Registration:** Clinicaltrial.gov identifier: NCT02545738 and NCT03449654

## Background

Type 2 diabetes (T2D) is a chronic disease that can lead to comorbidities and excess mortality. The risk of cardiovascular (CV) disease (CVD) is increased with T2D, making CVD the leading cause of death among individuals with T2D (1,2). Ceramides are a class of lipids associated with lipotoxic signaling, inflammation and apoptosis (3). They are present in the circulation as part of low-density lipoprotein (LDL) and very low-density lipoprotein (VLDL) particles (4). Recent studies have identified a set of specific ceramides that are strongly associated with the risk of CVD (5–9). Specifically, Ceramide(d18:1/16:0), Ceramide(d18:1/18:0), Ceramide(d18:1/20:0), Ceramide(d18:1/22:0), Ceramide(d18:1/24:0) and Ceramide(d18:1/24:1), which will be referred to in their abbreviated names as C16 Cer, C18 Cer, C20 Cer, C22 Cer, C24 Cer, and C24:1 Cer, respectively. C16 Cer, C18 cer, and C24:1 Cer are associated with increased CVD risk (6,9), while C22 Cer and C24 Cer are associated with decreased CVD risk (7). C20 Cer and C22 Cer appear to be weaker CVD predictors as there have been conflicting reports (5,8).

Individuals with T2D typically have elevated levels of C16 Cer, C18 Cer, C20 Cer and C24:1 Cer (10–12). C16 Cer, C18 Cer, C20 Cer, C22 Cer and C24 Cer have been associated with insulin resistance (13–15) and C18 Cer, C20 Cer and C22 Cer have been linked to the risk of developing T2D (16–19). Improvement in insulin sensitivity following gastric bypass surgery has been correlated with reduction in C18 Cer and C22 Cer levels, and low baseline levels of these ceramides were found to predict a higher likelihood of diabetes remission (20,21). Studies in mice have shown that reducing these ceramides can reduce insulin resistance (22–24) and development of CVD (25,26).

We previously demonstrated, in the trial called Effect of Liraglutide on Vascular Inflammation in Type-2 Diabetes (LiraFlame26), that treatment with the glucagon-like peptide-1 receptor agonist (GLP-1RA) liraglutide (1.8 mg/d) downregulated several lipid species, including ceramides in individuals with T2D, using an untargeted lipidomic approach (27). Conversely, GLP-1RAs, including liraglutide, have been shown to reduce the risk of CV outcomes (28–31). A better understanding of liraglutide’s effect on ceramide dynamics could provide insight into the observed cardioprotective effects of GLP-1RAs. By reducing lipotoxic molecules, drug therapies for diabetes could facilitate managing diabetes and its complications. In this study, we conducted a post-hoc analysis of plasma ceramides from two randomized, double-blinded, placebo-controlled trials involving 27 and 102 individuals with T2D, respectively, to investigate the effect of liraglutide treatment on the CVD biomarkers C16 Cer, C18 Cer, C20 Cer, C22 Cer, C24 Cer, and C24:1 Cer.

## Methods

### Participants and study design

Plasma samples for this study were acquired from participants in two randomized clinical trials. The first trial, named Antiproteinuric Effects of Liraglutide Treatment (LirAlbu12), was a randomized double-blinded crossover clinical trial conducted at Steno Diabetes Center Copenhagen between 2015 and 2016. The trial, which aimed at studying the effect of liraglutide on albuminuria in T2D, has previously been described in detail (32). In brief, the trial included participants with T2D who had persistent albuminuria (≥30 mg/g in at least 2 of 3 urine samples at inclusion) and who received renin-angiotensin-system blocking therapy. The participants were randomly assigned to receive either 12 weeks of subcutaneous liraglutide (up to 1.8 mg/day) or placebo, in addition to standard care. After completing the first treatment regimen, the participants underwent a 4-week washout period before crossing over to the opposite treatment for the next 12 weeks. Plasma samples were collected before and after each treatment regimen and were analyzed for the 27 participants who completed the trial. The trial is registered at Clinicaltrial.gov under the identifier NCT02545738.

The second study included is a randomized double-blinded parallel clinical trial here called LiraFlame26. LiraFlame26 was carried out to evaluate anti-atherogenic effects of liraglutide in T2D, the protocol has previously been published (33). The trial was conducted at Steno Diabetes Center Copenhagen between 2017 and 2019 and recruited 102 participants with T2D who were randomized to either subcutaneous liraglutide (up to 1.8 mg/day) or placebo for 26 weeks. Plasma samples were collected at baseline and end of trial. LiraFlame26 was registered at Clinicaltrial.gov with the identifier NCT03449654.

Both trials were approved by regional ethics committee and the Danish Medicine Agency and followed the principles laid out by the Declaration of Helsinki and Good Clinical Practice. Participants provided written informed consent before their enrollment in the study.

### Lipid analyses

Lipids were extracted from the plasma samples obtained from the LirAlbu12 trial using a modified Folch procedure previously reported (34,35). The samples were prepared in random order before being analyzed, and pooled samples and blanks were included between every 12 samples for quality control. A set calibration curve samples were analyzed at the start and in the end of the analytical run, using a serial dilution ranging between 0.25 – 200 μg/ml of the pure reference standards C16 Cer, C18 Cer and C24 Cer. The extracted samples were analyzed using an Infinity II ultra-high-performance liquid chromatography system coupled with an Agilent 6550 quadrupole time-of-flight mass spectrometry from Agilent Technologies.

The raw mass spectrometry data from a previously published work of LiraFlame26 (27) were reanalyzed for this work, in parallel with and in the same manner as LirAlbu12. Preprocessing of the mass spectrometry data was carried out using MZmine2 v.2.28 (36), targeting the six ceramides of interest in positive ionization mode: C16 Cer, C18 Cer, C20 Cer, C22 Cer, C24 Cer and C24:1 Cer. The ceramides produced a water loss adduct [M-H2O+H]^+^, used as the quantifier ion, and a protonated adduct [M+H]^+^, used as a qualifier ion (37). The ceramide peaks in LirAlbu12 were normalized to a pure exogenous internal standard, Ceramide(d18:1/17:0) (C17 Cer), which was spiked into all samples at the same concentration. Calibration curves were constructed by linearly fitting the peak areas of the calibration curve standards, normalized by the internal standard, against their respective concentrations. Absolute quantification in the LirAlbu12 trial was carried out by fitting each ceramide to the closest standard curve. For C20 Cer, this meant fitting to the C18 Cer standard curve, while the C24 Cer standard curve was used to calculate the concentrations of C22 Cer and C24:1 Cer. In contrast, no calibration curves were available for the LiraFlame26 trial, “amount” signifies ceramide peak area normalized to the exogenous standard, C17 Cer, absolute concentrations were not obtained. Outliers, defined as measures more than 3 standard deviations away from the median, were truncated.

### Statistical Analyses

Baseline characteristics, presented as mean (SD) or n (%), were compared between the liraglutide and placebo groups using the chi-square test for categorical variables and ANOVA for continuous variables. These results were compiled into a table using the tableone package in R (38). To assess the changes in ceramide levels after each treatment, linear mixed models were built for each ceramide level as a function of treatment and time considering random effects between participants (27), using the lme4, lmerTest and ggeffects packages (39–41). Changes were visualized with boxplots depicting ceramide amount before and after each treatment regimen using the ggplot2 and ggpubr packages (42,43). The ceramide changes from the linear mixed models were not adjusted. To assess whether the changes in ceramide levels were indirectly influenced by changes in other clinical variables affected by liraglutide treatment, linear regression models and causal mediation analysis from the R-package “mediation” was applied (44). Specifically, body weight, HbA1c levels, and urine albumin excretion rate (UAER) was tested for mediation effects.

The LirAlbu12 study was designed as a crossover study, which allowed each participant to be their own control, thus increasing statistical confidence. Ceramide changes of LirAlbu12 were calculated with paired t-test of the endpoints (32). Correlation between the ceramides and possible confounders were explored with the following variables: HbA_1c_, weight, systolic blood pressure, age, diabetes duration, total cholesterol, LDL cholesterol, triglyceride, UAER, and estimated glomerular filtration rate (eGFR). Pearson correlation coefficients were calculated and the correlations were visualized using the Hmisc (45) and ggcorrplot packages (46). A two-sided p-value < 0.05 was considered statistically significant. The final processing, statistics and visualizations was carried out in R v.4.2.0 (47), the code is available on Github: https://github.com/Asger-W/Liraglutide-Ceramides.

## Results

### Baseline characteristics

The baseline characteristics of the participants are summarized in Table 1. LirAlbu12 included 27 participants with T2D and albuminuria, with a mean (SD) age of 65.3 (7.3) years, diabetes duration of 14.8 (7.1) years and 18.5% (n = 5) women. LiraFlame26 involved 102 individuals with T2D randomized to either liraglutide or placebo treatment, with a mean (SD) age of 66.4 (8.2) years, diabetes duration of 13 (8.7) years and 15.7% (n = 16) women. At baseline LiraFlame26 participants in the liraglutide group had by chance higher triglyceride levels compared to the placebo group 2.07 (1.19) vs. 1.56 (0.78) mmol/L (p = 0.013), but the studies were otherwise balanced. The baseline characteristics of the two trials were overall comparable.

**Table 1.** Baseline characteristics.

| **Baseline characteristics LirAlbu12** |  |  |  |  |
| --- | --- | --- | --- | --- |
|  | **Start of trial**<br>**Total** | **Before**<br>**Liraglutide** | **Before**<br>**Placebo** | **P-value** |
| n | 27 | 27 | 27 |  |
| Age (years) | 65.3 (7.3) | 65.3 (7.3) | 65.3 (7.3) | 1 |
| Woman | 5 (18.5 %) | 5 (18.5 %) | 5 (18.5 %) | 1 |
| Weight (kg) | 99.5 (18.7) | 100 (18.5) | 99.5 (17.8) | 0.923 |
| Diabetes duration (years) | 14.8 (7.1) | 14.8 (7.1) | 14.8 (7.1) | 1 |
| HbA<sub>1c</sub> (mmol/mol) | 62 (10.7) | 62.6 (11.6) | 59.6 (12.1) | 0.356 |
| HbA<sub>1c</sub> (%) | 7.8 (1) | 7.9 (1.1) | 7.6 (1.1) | 0.356 |
| Total cholesterol (mmol/L) | 3.8 (0.9) | 3.8 (1) | 3.8 (0.8) | 0.886 |
| LDL (mmol/L) | 1.88 (0.59) | 1.82 (0.50) | 1.86 (0.61) | 0.768 |
| Triglyceride (mmol/L) | 2.20 (1.79) | 1.99 (1.31) | 2.26 (1.70) | 0.518 |
| Systolic blood pressure (mm Hg) | 136 (18) | 137 (16) | 133 (15) | 0.355 |
| eGFR (mL/min/1.73m<sup>2</sup>) | 75 (23) | 75 (23) | 73 (23) | 0.783 |
| UAER (mg/d) | 8 [7 – 13] | 9 [7 – 14] | 8 [7 – 12] | 0.967 |
| **Baseline characteristics LiraFlame26** |  |  |  |  |
|  | **Total** | **Liraglutide** | **Placebo** | **P-value** |
| n | 102 | 51 | 51 |  |
| Age (years) | 66.4 (8.2) | 65.9 (8.6) | 66.9 (7.8) | 0.556 |
| Women | 16 (15.7 %) | 6 (11.8 %) | 10 (19.6 %) | 0.414 |
| Weight (kg) | 91.2 (17.3) | 94.5 (19.9) | 87.9 (13.6) | 0.055 |
| Diabetes duration (years) | 13 (8.7) | 13.3 (9.1) | 12.6 (8.3) | 0.657 |
| HbA<sub>1c</sub> (mmol/mol) | 58.4 (10.1) | 58.7 (9.6) | 58 (10.6) | 0.725 |
| HbA<sub>1c</sub> (%) | 7.5 (0.9) | 7.5 (0.9) | 7.5 (1) | 0.725 |
| Total cholesterol (mmol/L) | 4.1 (0.8) | 4.1 (0.8) | 4.1 (0.8) | 0.855 |
| LDL (mmol/L) | 2.10 (0.67) | 2.05 (0.72) | 2.15 (0.62) | 0.476 |
| Triglyceride (mmol/L) | 1.81 (1.03) | 2.07 (1.19) | 1.56 (0.78) | 0.013 |
| Systolic blood pressure (mm Hg) | 135 (17) | 133 (14) | 137 (20) | 0.253 |
| eGFR (mL/min/1.73m<sup>2</sup>) | 83 (16) | 83 (18) | 84 (15) | 0.746 |
| UAER (mg/d) | 2 [2 – 3] | 3 [2 – 4] | 2 [2 – 3] | 0.201 |

Mean (SD), n (%) or median [IQR], groupwise comparison between liraglutide and placebo was carried out using chi square test for categorical variables and ANOVA for continuous variables. LDL: Low-density lipoprotein, eGFR: Estimated glomerular filtration rate, UAER: Urinary albumin excretion rate.

### Ceramides reduced by liraglutide treatment

Changes in ceramide levels were investigated using linear mixed models allowing for random effect between individuals. The ceramide amount was measured as the ratio of ceramide intensity divided by the intensity of pure standard (peak area of ceramide / peak area ceramide standard). At the end of LiraFlame26 the liraglutide group had significantly lower levels of C16 Cer and C24:1 Cer compared to the placebo group. The estimated differences (SD) were -2.556e-04 (1.260e-04) and -1.342e-02 (6.680e-03) for C16 Cer and C24:1 Cer respectively, with p-values of 0.045 and 0.047 (Table 2). Specifically, the mean C16 Cer level was 9.5% lower in the liraglutide group after treatment compared to the placebo group, and the C24:1 Cer level was 18.4% lower. Although the other investigated ceramides were not significantly altered by liraglutide treatment compared to placebo, they showed a trend towards lower levels by intervention, except for C20 Cer which showed a small non-significant increase after liraglutide treatment compared to placebo.

**Table 2.** Linear mixed models.

|  | LMM LirAlbu12 |  |  | LMM LiraFlame26 |  |  |
| --- | --- | --- | --- | --- | --- | --- |
|  | Coefficient (µg/ml) | Standard Error | P-value | Coefficient | Standard Error | P-value |
| **C16 Cer** | -0.033 | 0.028 | 0.240 | -2.556e-04 | 1.260e-04 | 0.045* |
| **C18 Cer** | -0.015 | 0.033 | 0.639 | -6.000e-04 | 3.459e-04 | 0.086 |
| **C20 Cer** | -0.031 | 0.053 | 0.559 | 2.953e-05 | 2.629e-05 | 0.264 |
| **C22 Cer** | -0.239 | 0.252 | 0.344 | -1.272e-04 | 1.690e-04 | 0.454 |
| **C24 Cer** | -1.006 | 1.075 | 0.352 | -3.108e-02 | 1.929e-02 | 0.110 |
| **C24:1 Cer** | 0.089 | 0.326 | 0.786 | -1.342e-02 | 6.680e-03 | 0.047* |

Linear mixed models were constructed for each ceramide in both trials with the following formular:
*Ceramide ~ Treatment+Time + Treatment:Time + (1/Participant ID)*. \* Indicate p-value <0.05.

The ceramides changes in the LirAlbu12 trial showed a similar trend to the LiraFlame26, where the overall ceramide levels decreased after liraglutide treatment and slightly increased after the placebo treatment (Figure 1). However, these changes were not statistically significant in the linear mixed modeling (Table 2). Paired t-test of end-to-end differences in the LirAlbu12 trial showed that liraglutide treatment led to significantly lower levels of C16 Cer and C24 Cer compared to placebo. After liraglutide treatment, the mean (SD) C16 Cer levels were 0.732 (0.149) μg/ml, and after placebo treatment, they were 0.772 (0.167) μg/ml. The mean (95% CI) C16 Cer level difference between liraglutide and placebo was -0.039 (−0.075; -0.002) μg/ml, which corresponds to a difference of -4.6% (p = 0.039). Similarly, for C24 Cer, the mean (SD) amount after liraglutide treatment was 7.984 (4.329) μg/ml, and after placebo treatment, it was 9.499 (5.445) μg/ml. The mean (95% CI) C24 Cer level difference between liraglutide and placebo was -1.729 (−3.418; -0.04) μg/ml, which corresponds to a difference of -7.5% (Table 3).

**Table 3.**
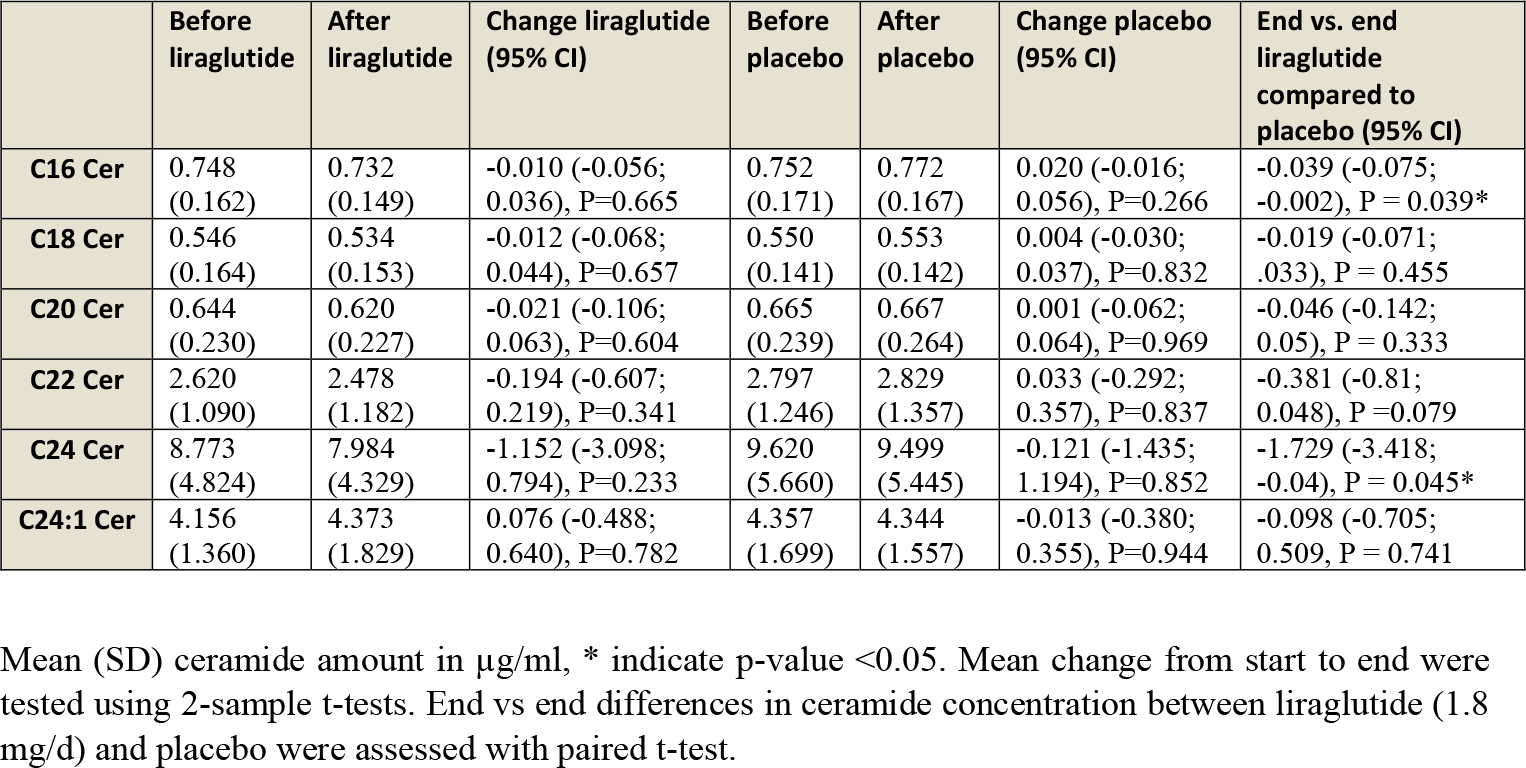
Ceramide measures by treatment group in LirAlbu12.

|  | Before<br>liraglutide | After<br>liraglutide | Change liraglutide<br>(95% CI) | Before<br>placebo | After<br>placebo | Change placebo<br>(95% CI) | End vs. end<br>liraglutide<br>compared to<br>placebo (95% CI) |
| --- | --- | --- | --- | --- | --- | --- | --- |
| **C16 Cer** | 0.748<br>(0.162) | 0.732<br>(0.149) | -0.010 (-0.056;<br>0.036), $P=0.665$ | 0.752<br>(0.171) | 0.772<br>(0.167) | 0.020 (-0.016;<br>0.056), $P=0.266$ | -0.039 (-0.075;<br>-0.002), $P = 0.039^*$ |
| **C18 Cer** | 0.546<br>(0.164) | 0.534<br>(0.153) | -0.012 (-0.068;<br>0.044), $P=0.657$ | 0.550<br>(0.141) | 0.553<br>(0.142) | 0.004 (-0.030;<br>0.037), $P=0.832$ | -0.019 (-0.071;<br>.033), $P = 0.455$ |
| **C20 Cer** | 0.644<br>(0.230) | 0.620<br>(0.227) | -0.021 (-0.106;<br>0.063), $P=0.604$ | 0.665<br>(0.239) | 0.667<br>(0.264) | 0.001 (-0.062;<br>0.064), $P=0.969$ | -0.046 (-0.142;<br>0.05), $P = 0.333$ |
| **C22 Cer** | 2.620<br>(1.090) | 2.478<br>(1.182) | -0.194 (-0.607;<br>0.219), $P=0.341$ | 2.797<br>(1.246) | 2.829<br>(1.357) | 0.033 (-0.292;<br>0.357), $P=0.837$ | -0.381 (-0.81;<br>0.048), $P = 0.079$ |
| **C24 Cer** | 8.773<br>(4.824) | 7.984<br>(4.329) | -1.152 (-3.098;<br>0.794), $P=0.233$ | 9.620<br>(5.660) | 9.499<br>(5.445) | -0.121 (-1.435;<br>1.194), $P=0.852$ | -1.729 (-3.418;<br>-0.04), $P = 0.045^*$ |
| **C24:1 Cer** | 4.156<br>(1.360) | 4.373<br>(1.829) | 0.076 (-0.488;<br>0.640), $P=0.782$ | 4.357<br>(1.699) | 4.344<br>(1.557) | -0.013 (-0.380;<br>0.355), $P=0.944$ | -0.098 (-0.705;<br>0.509), $P = 0.741$ |

Mean (SD) ceramide amount in $\mu\text{g/ml}$ , \* indicate $p\text{-value} < 0.05$ . Mean change from start to end were tested using 2-sample t-tests. End vs end differences in ceramide concentration between liraglutide (1.8 mg/d) and placebo were assessed with paired t-test.

**Figure 1.**
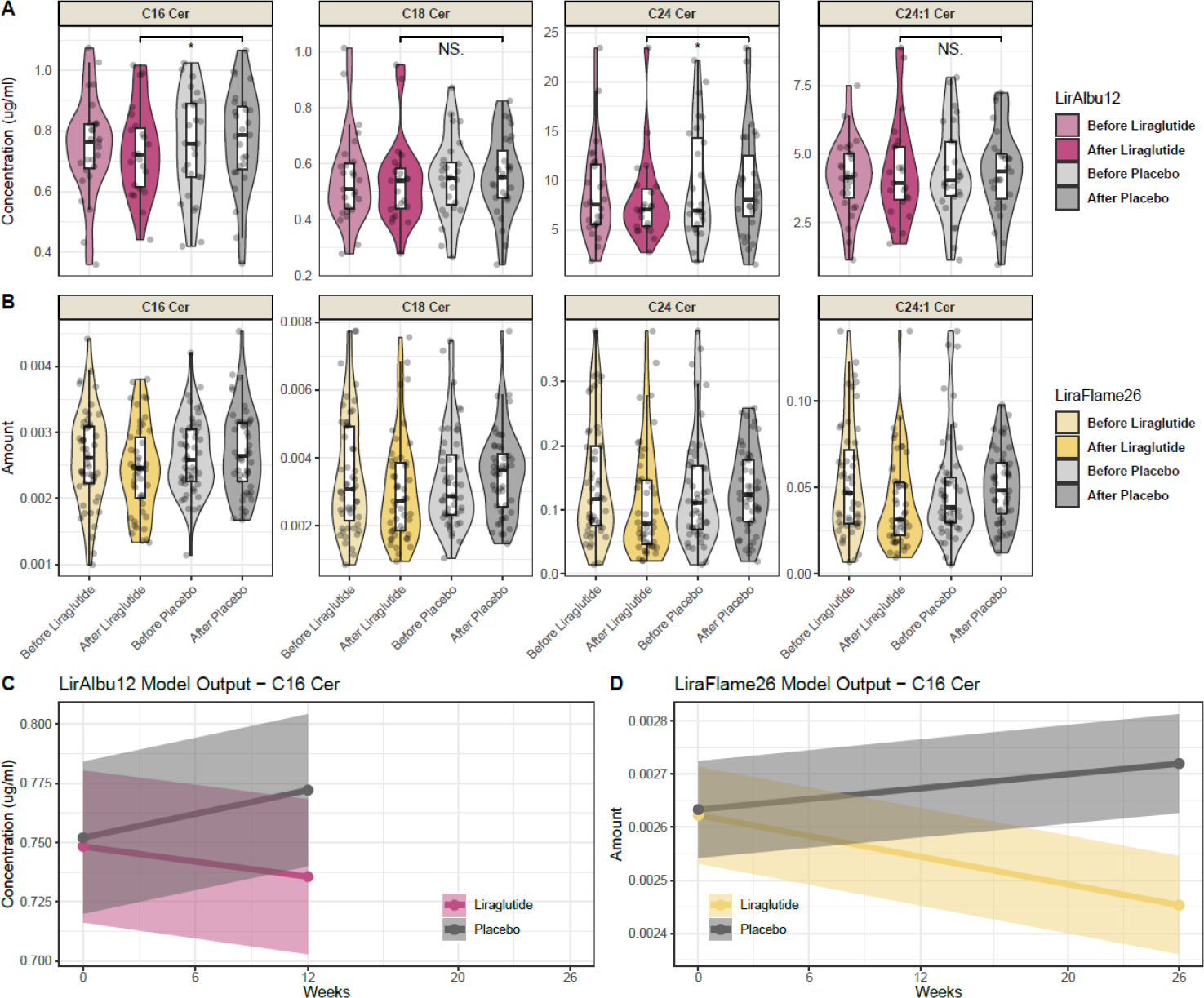
Ceramide distribution at different time points. Ceramide distribution at different time points. Boxplots of ceramides before and after liraglutide and placebo in (A) LirAlbu12, with * indicating paired t-test p-value < 0.05, and (B) LiraFlame26. Line graphs of C16 Cer amount per week over the two trials, modeled as linear mixed models: *Ceramide ~ Treatment+Time + Treatment:Time + (1*|*Participant ID)*, for (C) LirAlbu12 and (D) LiraFlame26. The ceramide amount was measured as the ratio of ceramide intensity divided by the intensity of pure standard (peak area of ceramide / peak area ceramide standard).

### Ceramides in relation to weight and other outcomes

To understand the interplay between ceramide concentrations and outcome variables, their correlations were investigated. The ceramides were positively correlated with other lipid measures such as triglycerides and total cholesterol (correlation estimates between 0.19-0.55, p < 0.05), but not LDL cholesterol (Figure 2). No ceramide showed significant correlation to blood pressure or weight loss.

**Figure 2.**
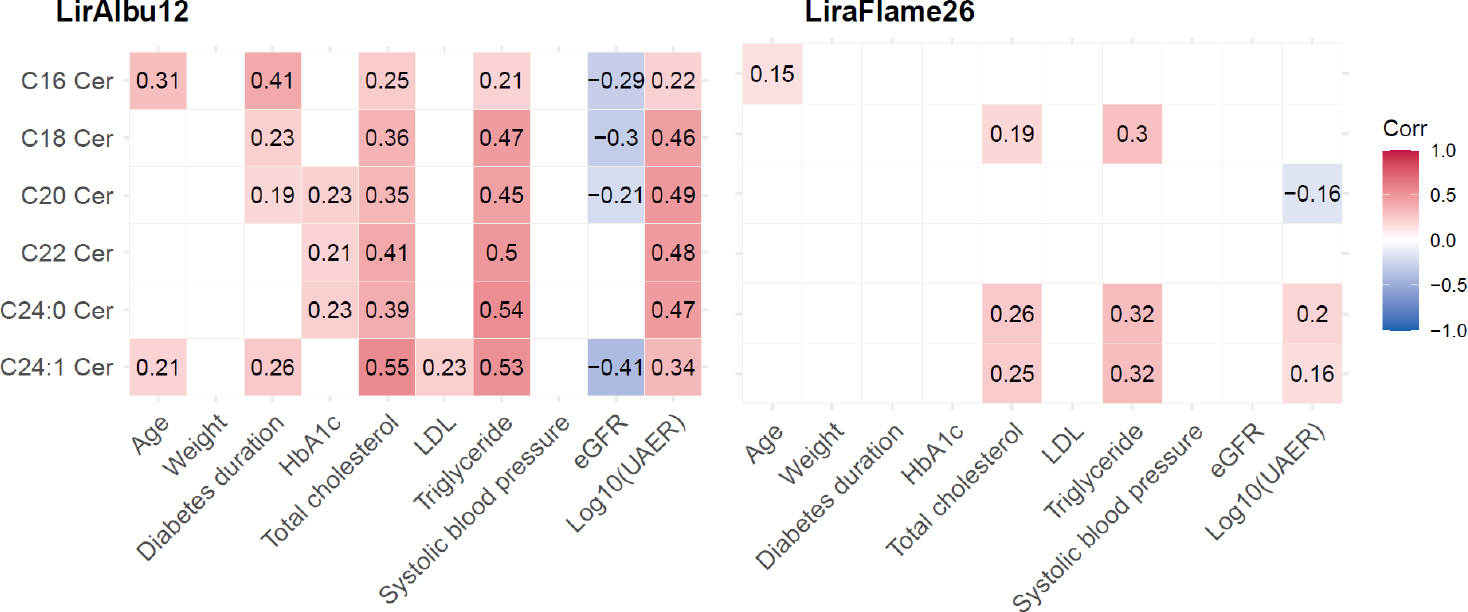
Ceramide correlation matrix. Pearson’s correlation between ceramides and possible confounders, numbers indicate estimates, only correlation with a p-value < 0.05 is shown. LDL: Low-density lipoprotein, eGFR: Estimated glomerular filtration rate, UAER: Urinary albumin excretion rate.

Weight loss did not mediate changes in ceramides levels in any of the trials, with p > 0.05 for the average causal mediation effect (ACME) (Supplementary Table 1 and 2). Thus, body weight loss is not a requirement for lowering ceramides with liraglutide.

In the LirAlbu12 trial C20 Cer, C22 Cer and C24 Cer all showed significant correlation to HbA_1c_ with estimates (95% CI) 0.23 (0.042; 0.407), 0.21 (0.02; 0.388), 0.23 (0.035; 0.401) and p = 0.017, 0.031, 0.021 respectively. However, no such correlation between ceramides and HbA1c was found in the LiraFlame26 trial.

Mediation analysis showed divergent results in both trials for HbA1c and UAER. The LirAlbu12 trial suggested that changes in HbA1c may be influencing some of the changes seen in C16 Cer, C22 Cer, and C24 Cer, mediation, assessed by ACME, showed p-values of 0.034, 0.028, and 0.008, respectively. In contrast no indication of mediation for HbA1c was found in the LiraFlame26 trial.

Similarly, for UAER levels, the results were inconclusive: In the LiraFlame26 trial, C24 Cer and C24:1 Cer show positive correlation and C20 Cer show a negative correlation to UAER. In contrast, all investigated ceramides in the LirAlbu12 trial showed a significant positive correlation to UAER (correlation estimates between 0.22-0.49), one the main outcomes of the LirAlbu12 trial. Furthermore, changes in UAER appeared to mediate the changes of C20 Cer, C22 Cer, C24 Cer and C24:1 Cer, but not C16 Cer, mediation, assessed by ACME, resulted in p-values of 0.03, 0.016, 0.008, 0.026 and 0.872 respectively in the LirAlbu12 trial. No indication of UAER mediation was found in the LiraFlame26 trial with all ACME p > 0.05 (Supplementary Table 2).

## Discussion

This study examined the impact of liraglutide treatment on ceramides that have previously been shown associated with CV risk and all-cause mortality. The results showed that individuals treated with liraglutide had significantly lower levels of C16 Cer compared to placebo treated after both 12 and 26-weeks of treatment. While not all the ceramides investigated reached changes that were statistical significance, there was a consistent trend of decreasing ceramide levels following liraglutide treatment across two independent clinical trials.

### Therapy for ceramide reduction

Our findings correspond well with previous studies on the effects of liraglutide and other GLP-1RAs on ceramides (12,48–50). One study, on people with obesity (n = 32), found that after one year of follow-up, those treated with liraglutide (1.2 mg/d) maintained a stable level of C16 Cer, whereas those without liraglutide treatment had an increase in the level of C16 Cer (49). Jendle *et al*. reported a decrease in ceramides after 18 weeks of treatment with liraglutide (1.8 mg/d) and the sulfonylurea glimepiride (4 mg/d) respectively, in individuals with T2D (n = 62), ceramides decreased more with liraglutide treatment than with glimepiride (50). In the phase 2 trial for tirzepatide, a dual-agonist activating both the GLP-1 and the glucose-dependent insulinotropic polypeptide receptors, people (n = 314) with T2D were randomized to placebo or tirzepatide treatment for 26 weeks. The levels of C22 Cer and C24 Cer were lower in people treated with tirzepatide (15 mg/w) in comparison to the placebo group (48). Zhang *et al*. investigated the GLP-1RA exenatide and reported unchanged ceremide levels for the six ceramides in people (n = 35) with T2D after 12 weeks of exenatide treatment (20 μg/d) (12). Taken together, these findings suggest that lowering of ceramide is a likely effect of GLP-1RAs treatments, but to a varying extent. Furthermore, several studies reported improvements in lipoprotein profiles following treatment with liraglutide (51–54), indicating that its effects on the lipidome may not be limited to ceramides. The lipid modulating effects of liraglutide and other GLP-1RAs are worth considering when selecting treatments for individuals with T2D, particularly for those with highest CVD risk.

Unlike other ceramides, C24 Cer is in some studies associated with CVD protection, meaning that lower levels of C24 Cer are associated with increased CVD risk (5,8). Our findings suggest that liraglutide lowers C24 Cer, together with the lipotoxic ceramides. Further studies are needed to establish protective roles of ceramides and if decrease in these species outweighs the benefits of lowering lipotoxic ceramides.

### Ceramide in relation to clinical outcomes

The LiraFlame26 trial showed fewer correlations between ceramides and other risk factors compared to the LirAlbu12 trial. These differences could be because the number of participants was smaller in the LirAlbu12 trial or because of the crossover design, led to lower ceramide variation. Interestingly, weight loss did not mediate the reduction of ceramide levels in our studies. This is in agreement with Akawi et al. who observed suppression of C16 Cer following one year liraglutide treatment (1.2 mg/d) compared to placebo (n = 32), but did not observe changes in BMI (49). Other studies have observed a simultaneous reduction of ceramide levels and weight, but have not investigated, whether the weight loss mediates the reduction in ceramide levels (20,55,56).

It is worth noting that, unlike the LiraFlame26 trial, the LirAlbu12 trial had an inclusion criterion on albuminuria. Mediation analysis suggested that some of the observed effect of ceramides reduction in the LirAlbu12 trial could be mediated by a reduction in HbA_1C_ and UAER. The ceramides investigated in the present study have previously been linked to kidney disease. A study by Liu *et al*. found that C16 Cer levels were significantly higher in people with overt diabetic kidney disease (57). Similarly, Mantovani *et al*. found increased levels of all six ceramides, investigated in this paper, in people with chronic kidney disease compared to those without chronic kidney disease (58). Comparable findings have been made in a mouse model (59). As lower UAER and lower ceramide levels both are linked to improved kidney health, a correlation between UAER and ceramide levels could be expected.

In our previous work, we found that liraglutide treatment resulted in a lower activity of stearoyl-CoA desaturase-1 (SCD1) (60). SCD1 is a lipogenic enzyme responsible for conversion of saturated fatty acids to monounsaturated fatty acids. One study found that SCD1 deficient mice had approximately 40% reduction in ceramides (61). This could potentially help explain the observed reduction in ceramide levels in response to liraglutide treatment. Here we demonstrated that liraglutide consistently lowered C16 Cer levels in individuals with T2D, compared to placebo, despite the effect of ceramide change was relatively modest and the interindividual differences showed high variation.

### Strengths and limitations

A strength of the present study is that it combines the effects observed in two independent trials one with a crossover and one with a parallel trial design. Another strength is analyses of the LirAlbu12 trial with measurement of the absolute concentrations of six pre-specified ceramides, making the findings translatable between laboratories and populations.

A limitation for this post-hoc study is that it was not designed with ceramides as an outcome. More studies are needed to determine if longer treatment with liraglutide can significantly reduce the concentrations of ceramides associated to CVD. Additionally, we did not control for diet, exercise, or medication changes, all of which could affect ceramide levels.

## Conclusions

We have investigated the impact of liraglutide treatment on ceramides in two clinical trials including individuals with T2D, as the ceramides a known to be associated with CVD risk. We found that C16 Cer was significantly reduced in both trials with liraglutide treatment compared to placebo and all ceramide levels were consistently reduced, however not to significance in the timespan of 26 weeks. These results suggest that liraglutide treatment can be used as a ceramide lowering intervention which could potentially have significant implications in precision medicine initiatives for CV risk. However, further research is needed to determine if lowering of ceramides translates to decreased CV risk.

## Data Availability

The dataset analyzed here is not publicly available, for the privacy of the participants, in compliance with EU and Danish data protection law. The data can be accessed upon reasonable request; relevant legal permission from the data protection agency is required. Data access request should be directed to PR.

https://github.com/Asger-W/Liraglutide-Ceramides

## List of abbreviations

ADE: Average direct effect
ACME: average causal mediation effect
Cer: Ceramide
CVD: Cardiovascular disease
C16 Cer: ceramide(d18:1/16:0)
C18 Cer: ceramide(d18:1/18:0)
C17 Cer: ceramide(d18:1/17:0)
C20 Cer: ceramide(d18:1/20:0)
C22 Cer: ceramide(d18:1/22:0)
C24 Cer: ceramide(d18:1/24:0)
C24:1 Cer: ceramide(d18:1/24:1)
eGFR: Estimated glomerular filtration rate
HbA1c: Hemoglobin A1C
LDL: Low-density lipids
RA: Receptor agonist
SCD1: stearoyl-CoA desaturase-1
T2D: Type 2 Diabetes
UAER: Urinary albumin excretion rate

## Declarations

### Availability of data and materials

The code used for data analysis is available on github: https://github.com/Asger-W/Liraglutide-Ceramides.

### Competing interests

The authors declare no conflict of interest compromising the integrity of this work.

#### Disclosures outside this work

EZ and BJvS are now employees at Novo Nordisk and have shares in Novo Nordisk. RR has served as consultant for Novo Nordisk and has shares in Novo Nordisk. AK has served on advisory boards or as consultant for Novo Nordisk, Curium, Clarity Pharmaceuticals, IPSEN, and Siemens Healthineers. TH own shares in Novo Nordisk. TV has served on scientific advisory panels, been part of speaker’s bureaus, served as a consultant to and/or received research support from Amgen, AstraZeneca, Boehringer Ingelheim, Eli Lilly, Gilead, GSK, Mundipharma, Novo Nordisk, Sanofi and Sun Pharmaceuticals. PR has received honoraria for consultancy to Steno Diabetes Center Copenhagen from Astellas, Astra Zeneca, Boehringer Ingelheim, Bayer, Merck, Gilead, Novo Nordisk, Sanofi Aventis. CLQ has served on scientific advisory panels and/or received research support from Pfizer, Novo Nordisk and other companies via the IMI funding scheme.

### Funding

Liraflame was funded by Novo Nordisk A/S and Skibsreder Per Henriksen, R. og hustrus fund. Steno Diabetes Center Copenhagen and Department of Clinical Physiology, Nuclear Medicine & PET, Rigshospitalet & Cluster for Molecular Imaging, University of Copenhagen, Denmark have provided internal funding (ERC Advanced Grant no. 670261; Research Foundation of Rigshospitalet; Research Council of the Capital Region of Denmark; Lundbeck Foundation; Novo Nordisk Foundation; The John and Birthe Meyer Foundation).

### Authors’ contributions

AW, HV, PR and CLQ contributed to the conceptualization and interpretation of this study, BJH conducted the LirAlbu12 study, EZ, VRC, TH, RR and AK carried out the LiraFlame26 study and provided material and clinical data for this study. AW and AZ performed mass spectrometry analysis. AW, JX and TS contributed to preprocessing of the mass spectrometry data. AW performed data analysis and drafted the manuscript. All authors approved the final version of the manuscript. AWR and CLQ are responsible for the integrity of the work as a whole.

## Notes

### Clinical Trial

ClinicalTrials.gov Identifier: NCT03449654 and NCT02545738

### Author Declarations

The protocol was approved by the local ethics committee at RegionH Denmark (H-16044546) and the Danish Medicines Agency (2016110109)

## References

1. Tancredi M, Rosengren A, Svensson A-M, Kosiborod M, Pivodic A, Gudbjörnsdottir S et al. Excess Mortality among Persons with Type 2 Diabetes. N Engl J Med. 2015;373(18):1720–32. doi: 10.1056/nejmoa1504347

2. Baena-Díez JM, Peñafiel J, Subirana I, Ramos R, Elosua R, Marín-Ibañez A et al. Risk of cause-specific death in individuals with diabetes: A competing risks analysis. Diabetes Care. 2016;39(11):1987–95. doi: 10.2337/dc16-0614

3. Chaurasia B, & Summers SA. Ceramides in Metabolism: Key Lipotoxic Players. Annu Rev Physiol. 2021;83:303–30. doi: 10.1146/annurev-physiol-031620-093815

4. Kotronen A, Velagapudi VR, & Yetukuri L. Serum saturated fatty acids containing triacylglycerols are better markers of insulin resistance than total serum triacylglycerol concentrations. Diabetologia. 2009;684–90. doi: 10.1007/s00125-009-1282-2

5. Tarasov K, Ekroos K, Suoniemi M, Kauhanen D, Sylvänne T, Hurme R et al. Molecular lipids identify cardiovascular risk and are efficiently lowered by simvastatin and PCSK9 deficiency. J Clin Endocrinol Metab. 2014;99(1):45–52. doi: 10.1210/jc.2013-2559

6. Havulinna AS, Sysi-Aho M, Hilvo M, Kauhanen D, Hurme R, Ekroos K et al. Circulating Ceramides Predict Cardiovascular Outcomes in the Population-Based FINRISK 2002 Cohort. Arterioscler Thromb Vasc Biol. 2016;36(12):2424–30. doi: 10.1161/ATVBAHA.116.307497

7. Peterson LR, Xanthakis V, Duncan MS, Gross S, Friedrich N, Völzke H et al. Ceramide remodeling and risk of cardiovascular events and mortality. J Am Heart Assoc. 2018;7(10). doi: 10.1161/JAHA.117.007931

8. Lemaitre RN, Jensen PN, Hoofnagle A, Mcknight B, Fretts AM, King IB et al. Plasma Ceramides and Sphingomyelins in Relation to Heart Failure Risk: The Cardiovascular Health Study. Circ Hear Fail. 2019;12(7):1–8. doi: 10.1161/CIRCHEARTFAILURE.118.005708

9. Hilvo M, Meikle PJ, Pedersen ER, Tell GS, Dhar I, Brenner H et al. Development and validation of a ceramide- And phospholipid-based cardiovascular risk estimation score for coronary artery disease patients. Eur Heart J. 2020;41(3):371–80. doi: 10.1093/eurheartj/ehz387

10. Haus JM, Kashyap SR, Kasumov T, Zhang R, Kelly KR, Defronzo RA et al. Plasma ceramides are elevated in obese subjects with type 2 diabetes and correlate with the severity of insulin resistance. Diabetes. 2009;58(2):337–43. doi: 10.2337/db08-1228

11. Bergman BC, Brozinick JT, Strauss A, Bacon S, Kerege A, Bui HH et al. Serum sphingolipids: Relationships to insulin sensitivity and changes with exercise in humans. Am J Physiol - Endocrinol Metab. 2015;309(4):E398–408. doi: 10.1152/ajpendo.00134.2015

12. Zhang L, Hu Y, An Y, Wang Q, Liu J, & Wang G. The Changes of Lipidomic Profiles Reveal Therapeutic Effects of Exenatide in Patients With Type 2 Diabetes. Front Endocrinol (Lausanne). 2022;13(March):1–10. doi: 10.3389/fendo.2022.677202

13. Błachnio-Zabielska AU, Baranowski M, Hirnle T, Zabielski P, Lewczuk A, Dmitruk I et al. Increased bioactive lipids content in human subcutaneous and epicardial fat tissue correlates with insulin resistance. Lipids. 2012;47(12):1131–41. doi: 10.1007/s11745-012-3722-x

14. Luukkonen PK, Zhou Y, Sädevirta S, Leivonen M, Arola J, Orešič M et al. Hepatic ceramides dissociate steatosis and insulin resistance in patients with non-alcoholic fatty liver disease. J Hepatol. 2016;64(5):1167–75. doi: 10.1016/j.jhep.2016.01.002

15. Lemaitre RN, Yu C, Hoofnagle A, Hari N, Jensen PN, Fretts AM et al. Circulating sphingolipids, insulin, HOMA-IR, and HOMA-B: The Strong heart family study. Diabetes. 2018;67(8):1663–72. doi: 10.2337/db17-1449

16. Wigger L, Cruciani-Guglielmacci C, Nicolas A, Denom J, Fernandez N, Fumeron F et al. Plasma Dihydroceramides Are Diabetes Susceptibility Biomarker Candidates in Mice and Humans. Cell Rep. 2017;18(9):2269–79. doi: 10.1016/j.celrep.2017.02.019

17. Hilvo M, Salonurmi T, Havulinna AS, Kauhanen D, Pedersen ER, Tell GS et al. Ceramide stearic to palmitic acid ratio predicts incident diabetes. Diabetologia. 2018;61(6):1424–34. doi: 10.1007/s00125-018-4590-6

18. Chew WS, Torta F, Ji S, Choi H, Begum H, Sim X et al. Large-scale lipidomics identifies associations between plasma sphingolipids and T2DM incidence. JCI Insight. 2019;4(13):1–14. doi: 10.1172/jci.insight.126925

19. Fretts AM, Jensen PN, Hoofnagle A, McKnight B, Howard B V., Umans J et al. Plasma ceramide species are associated with diabetes risk in participants of the strong heart study. J Nutr. 2020;150(5):1214–22. doi: 10.1093/jn/nxz259

20. Huang H, Kasumov T, Gatmaitan P, Heneghan HM, Kashyap SR, Schauer PR et al. Gastric bypass surgery reduces plasma ceramide subspecies and improves insulin sensitivity in severely obese patients. Obesity [Internet]. 2011;19(11):2235–40. doi: 10.1038/oby.2011.107

21. Poss AM, Krick B, Maschek JA, Haaland B, Cox JE, Karra P et al. Following Roux-en-Y gastric bypass surgery, serum ceramides demarcate patients that will fail to achieve normoglycemia and diabetes remission. Med [Internet]. 2022;3(7):452–467.e4. doi: 10.1016/j.medj.2022.05.011

22. Holland WL, Brozinick JT, Wang LP, Hawkins ED, Sargent KM, Liu Y et al. Inhibition of Ceramide Synthesis Ameliorates Glucocorticoid-, Saturated-Fat-, and Obesity-Induced Insulin Resistance. Cell Metab. 2007;5(3):167–79. doi: 10.1016/j.cmet.2007.01.002

23. Ussher JR, Koves TR, Cadete VJJ, Zhang L, Jaswal JS, Swyrd SJ et al. Inhibition of de novo ceramide synthesis reverses diet-induced insulin resistance and enhances whole-body oxygen consumption. Diabetes. 2010;59(10):2453–64. doi: 10.2337/db09-1293

24. Raichur S, Brunner B, Bielohuby M, Hansen G, Pfenninger A, Wang B et al. The role of C16:0 ceramide in the development of obesity and type 2 diabetes: CerS6 inhibition as a novel therapeutic approach. Mol Metab [Internet]. 2019;21(January):36–50. doi: 10.1016/j.molmet.2018.12.008

25. Hojjati MR, Li Z, Zhou H, Tang S, Huan C, Ooi E et al. Effect of myriocin on plasma sphingolipid metabolism and atherosclerosis in apoE-deficient mice. J Biol Chem [Internet]. 2005;280(11):10284–9. doi: 10.1074/jbc.M412348200

26. Reforgiato MR, Milano G, Fabriàs G, Casas J, Gasco P, Paroni R et al. Inhibition of ceramide de novo synthesis as a postischemic strategy to reduce myocardial reperfusion injury. Basic Res Cardiol. 2016;111(2). doi: 10.1007/s00395-016-0533-x

27. Zobel EH, Wretlind A, Ripa RS, Curovic VR, von Scholten BJ, Suvitaival T et al. Ceramides and phospholipids are downregulated with liraglutide treatment: results from the LiraFlame randomized controlled trial. BMJ Open Diabetes Res Care. 2021;9(1):e002395. doi: 10.1136/bmjdrc-2021-002395

28. Marso SP, Daniels GH, Brown-Frandsen K, Kristensen P, Mann JFE, Nauck MA et al. Liraglutide and Cardiovascular Outcomes in Type 2 Diabetes. N Engl J Med. 2016;375(4):311–22. doi: 10.1056/NEJMoa1603827

29. Svanström H, Ueda P, Melbye M, Eliasson B, Svensson A, Franzén S et al. Use of liraglutide and risk of major cardiovascular events: a register-based cohort study in Denmark and Sweden. Lancet Diabetes Endocrinol. 2019;7(February 2019). doi: 10.1016/S2213-8587(18)30320-6

30. Kristensen SL, Rørth R, Jhund PS, Docherty KF, Sattar N, Preiss D et al. Cardiovascular, mortality, and kidney outcomes with GLP-1 receptor agonists in patients with type 2 diabetes: a systematic review and meta-analysis of cardiovascular outcome trials. Lancet Diabetes Endocrinol. 2019;7(October 2019). doi: 10.1016/S2213-8587(19)30249-9

31. Nathan DM, Lachin JM, Bebu I, Burch HB, Buse JB, Cherrington AL et al. Glycemia Reduction in Type 2 Diabetes - Microvascular and Cardiovascular Outcomes. N Engl J Med [Internet]. 2022;387(12):1075–88. doi: 10.1056/NEJMoa2200436

32. von Scholten BJ, Persson F, Rosenlund S, Hovind P, Faber J, Hansen TW et al. The effect of liraglutide on renal function: A randomized clinical trial. Diabetes, Obes Metab. 2017;19(2):239–47. doi: 10.1111/dom.12808

33. Ripa RS, Zobel EH, Scholten BJ von, Jensen JK, Binderup T, Diaz LJ et al. Effect of Liraglutide on Arterial Inflammation Assessed as [18F]FDG Uptake in Patients With Type 2 Diabetes: A Randomized, Double-Blind, Placebo-Controlled Trial. Circ Cardiovasc Imaging. 2021;14(July). doi: 10.1161/CIRCIMAGING.120.012174

34. Folch J, Lees M, & Stanley GHS. A Simple Method for the Isolation and Purification of Total Lipides from Animal Tissues. J Biol Chem. 1957;226(3):497–509.

35. Tofte N, Suvitaival T, Ahonen L, Winther SA, Theilade S, Frimodt-møller M et al. Lipidomic analysis reveals sphingomyelin and phosphatidylcholine species associated with renal impairment and all-cause mortality in type 1 diabetes. Sci Rep. 2019;1–10. doi: 10.1038/s41598-019-52916-w

36. Pluskal T, Castillo S, Villar-briones A, & Ore M. MZmine 2: Modular framework for processing, visualizing, and analyzing mass spectrometry-based molecular profile data. BMC Bioinformatics. 2010;11(395):1–11.

37. Kim M, Nevado-Holgado A, Whiley L, Snowden SG, Soininen H, Kloszewska I et al. Association between Plasma Ceramides and Phosphatidylcholines and Hippocampal Brain Volume in Late Onset Alzheimer’s Disease. J Alzheimers Dis. 2017;60(3):809–17. doi: 10.3233/JAD-160645

38. Yoshida K, Bartel A, Chipman JJ, Bohn J, McGowan LDa, Barrett M et al. R Package “ tableone”: Create “Table 1” to Describe Baseline Characteristics with or without Propensity Score Weights. 2022;

39. Bates D, Mächler M, Bolker BM, & Walker SC. Fitting linear mixed-effects models using lme4. J Stat Softw. 2015;67(1). doi: 10.18637/jss.v067.i01

40. Kuznetsova A, Brockhoff PB, & Christensen RHB. lmerTest Package: Tests in Linear Mixed Effects Models. J Stat Softw. 2017;82(13):1–26. doi: 10.18637/JSS.V082.I13

41. Lüdecke D. ggeffects: Tidy Data Frames of Marginal Effects from Regression Models. J Open Source Softw. 2018;3(26):772. doi: 10.21105/joss.00772

42. Wickham H. ggplot2: Elegant Graphics for Data Analysis. [Internet]. New York: Springer-Verlag New York; 2016.

43. Kassambara A. Ggpubr: “Ggplot2” Based Publication Ready Plots. 2021;

44. Tingley D, Yamamoto T, Hirose K, Keele L, & Imai K. mediation: R Package for Causal Mediation Analysis. J Stat Softw. 2014;59(5).

45. Harrell FE, & Dupont C. Package “Hmisc”: Harrell Miscellaneous. 2022;

46. Kassambara A, & Patil I. Package ‘ ggcorrplot ‘: Visualization of a Correlation Matrix using “ggplot2.” 2022;

47. R Core Team (2018). R: A language and environment for statistical computing. [Internet]. Vienna, Austria.: R Foundation for Statistical Computing; 2018.

48. Ruotolo G, Roth K., Milligan P., Lin Y, Wilson J., Pirro V et al. Effects of tirzepatide, a novel dual GIP and GLP-1 receptor agonist, on lipid profiling in patients with type 2 diabetes. Eur Heart J. 2020;41(Supplement_2):3056. doi: 10.1093/ehjci/ehaa946.3056

49. Akawi N, Checa A, Antonopoulos AS, Akoumianakis I, Daskalaki E, Kotanidis CP et al. Fat-Secreted Ceramides Regulate Vascular Redox State and Influence Outcomes in Patients With Cardiovascular Disease. J Am Coll Cardiol. 2021;77(20):2494–513. doi: 10.1016/j.jacc.2021.03.314

50. Jendle J, Hyötyläinen T, Orešič M, & Nyström T. Pharmacometabolomic profiles in type 2 diabetic subjects treated with liraglutide or glimepiride. Cardiovasc Diabetol [Internet]. 2021;20(1):1–12. doi: 10.1186/s12933-021-01431-2

51. Armstrong MJ, Hull D, Guo K, Barton D, Hazlehurst JM, Gathercole LL et al. Glucagon-like peptide 1 decreases lipotoxicity in non-alcoholic steatohepatitis. J Hepatol [Internet]. 2016;64(2):399–408. doi: 10.1016/j.jhep.2015.08.038

52. Engelbrechtsen L, Lundgren J, Wewer Albrechtsen NJ, Mahendran Y, Iepsen EW, Finocchietto P et al. Treatment with liraglutide may improve markers of CVD reflected by reduced levels of apoB. Obes Sci Pract. 2017;3(4):425–33. doi: 10.1002/osp4.133

53. Matikainen N, Söderlund S, Björnson E, Pietiläinen K, Hakkarainen A, Lundbom N et al. Liraglutide treatment improves postprandial lipid metabolism and cardiometabolic risk factors in humans with adequately controlled type 2 diabetes: A single-centre randomized controlled study. Diabetes, Obes Metab. 2019;21(1):84–94. doi: 10.1111/dom.13487

54. Verges B, Barros JPP de, Bouillet B, Baillot-rudoni S, Rouland A, Petit JM et al. Liraglutide Increases the Catabolism of Apolipoprotein B100 –Containing Lipoproteins in Patients With Type 2 Diabetes and Reduces Proprotein Convertase Subtilisin / Kexin Type 9 Expression. Diabetes Care. 2021;44(April):1027–37. doi: 10.2337/dc20-1843

55. Dubé JJ, Amati F, Toledo FGS, Stefanovic-Racic M, Rossi A, Coen P et al. Effects of weight loss and exercise on insulin resistance, and intramyocellular triacylglycerol, diacylglycerol and ceramide. Diabetologia. 2011;54(5):1147–56. doi: 10.1007/s00125-011-2065-0

56. Promrat K, Longato L, Wands JR, & De la Monte SM. Weight loss amelioration of nonalcoholic steatohepatitis linked to shifts in hepatic ceramide expression and serum ceramide levels. Hepatol Res. 2011;41(8):754–62. doi: 10.1111/j.1872-034X.2011.00815.x

57. Liu JJ, Ghosh S, Kovalik JP, Ching J, Choi HW, Tavintharan S et al. Profiling of Plasma Metabolites Suggests Altered Mitochondrial Fuel Usage and Remodeling of Sphingolipid Metabolism in Individuals With Type 2 Diabetes and Kidney Disease. Kidney Int Reports [Internet]. 2017;2(3):470–80. doi: 10.1016/j.ekir.2016.12.003

58. Mantovani A, Lunardi G, Bonapace S, Dugo C, Altomari A, Molon G et al. Association between increased plasma ceramides and chronic kidney disease in patients with and without ischemic heart disease. Diabetes Metab [Internet]. 2021;47(1):101152. doi: 10.1016/j.diabet.2020.03.003

59. Sas KM, Nair V, Pradeep Kayampilly JB, & Jharna Saha HZ. Targeted Lipidomic and Transcriptomic Analysis Identifies Dysregulated Renal Ceramide Metabolism in a Mouse Model of Diabetic Kidney Disease. J Proteomics Bioinform. 2015;s14(734). doi: 10.4172/jpb.s14-002

60. Wretlind A, Zobel EH, de Zawadzki A, Ripa RS, Curovic VR, von Scholten BJ et al. Liraglutide Lowers Palmitoleate Levels in Type 2 Diabetes. A Post Hoc Analysis of the LIRAFLAME Randomized Placebo-Controlled Trial. Front Clin Diabetes Healthc. 2022;3(March):1–8. doi: 10.3389/fcdhc.2022.856485

61. Dobrzyn A, Dobrzyn P, Lee S, Miyazaki M, Cohen P, Asilmaz E et al. Stearoyl-CoA desaturase-1 deficiency reduces ceramide synthesis by downregulating serine palmitoyltransferase and increasing B-oxidation in skeletal muscle. Am J Physiol Endocrinol Metab. 2005;53706:599–607. doi: 10.1152/ajpendo.00439.2004.

