## Supplementary table 1 and 2 for "Ceramides are decreased after liraglutide treatment in two randomized clinical trials"

### Supplementary table 1 – Mediation analysis of LirAlbu12

| | **Weight** | | | |
|---|---|---|---|---|
| **Ceramide** | **Total effect estimate** | **Total effect p-value** | **ACME estimate** | **ACME p-value** |
| C16 Cer | 0.025 (-0.03;0.079) | 0.372 | -0.004 (-0.032;0.021) | 0.718 |
| C18 Cer | 0.024 (-0.038;0.088) | 0.492 | 0.003 (-0.028;0.034) | 0.758 |
| C20 Cer | 0.037 (-0.053;0.141) | 0.45 | 0.002 (-0.048;0.047) | 0.912 |
| C22 Cer | 0.281 (-0.21;0.775) | 0.262 | 0.031 (-0.187;0.241) | 0.724 |
| C24 Cer | 1.272 (-0.762;3.59) | 0.238 | -0.238 (-1.312;0.687) | 0.716 |
| C24:1 Cer | -0.025 (-0.687;0.642) | 0.958 | 0.074 (-0.205;0.403) | 0.616 |

| | **HbA1c** | | | |
|---|---|---|---|---|
| **Ceramide** | **Total effect estimate** | **Total effect p-value** | **ACME estimate** | **ACME p-value** |
| C16 Cer | 0.03 (-0.027;0.083) | 0.318 | 0.039 (0.002;0.084) | 0.034 |
| C18 Cer | 0.016 (-0.047;0.073) | 0.664 | 0.054 (-0.037;0.132) | 0.274 |
| C20 Cer | 0.023 (-0.068;0.125) | 0.7 | 0.099 (-0.012;0.205) | 0.094 |
| C22 Cer | 0.227 (-0.244;0.755) | 0.374 | 0.553 (0.078;1.025) | 0.028 |
| C24 Cer | 1.031 (-0.971;3.167) | 0.32 | 3.019 (0.465;5.82) | 0.008 |
| C24:1 Cer | -0.089 (-0.732;0.569) | 0.762 | 0.422 (-0.191;0.988) | 0.22 |

| | **Log10MeanUAER** | | | |
|---|---|---|---|---|
| **Ceramide** | **Total effect estimate** | **Total effect p-value** | **ACME estimate** | **ACME p-value** |
| C16 Cer | 0.03 (-0.024;0.085) | 0.304 | 0.002 (-0.022;0.031) | 0.872 |
| C18 Cer | 0.016 (-0.051;0.081) | 0.612 | 0.01 (-0.009;0.043) | 0.35 |
| C20 Cer | 0.024 (-0.072;0.133) | 0.65 | 0.047 (0.003;0.127) | 0.03 |
| C22 Cer | 0.241 (-0.264;0.74) | 0.346 | 0.286 (0.035;0.688) | 0.016 |
| C24 Cer | 1.189 (-0.891;3.277) | 0.286 | 1.272 (0.236;2.673) | 0.008 |
| C24:1 Cer | -0.069 (-0.675;0.558) | 0.828 | 0.266 (0.022;0.739) | 0.026 |

Mediation effect was assessed by linear regression models with and without adjustment for the tested confounder the effect and significance were estimated by bootstrapping 1000 times using the mediator package in R. ACME:  Average casual mediation effect, UAER: Urinary albumin excretion rate.

**Supplementary table 2 – Mediation analysis of LiraFlame26**

| | **Weight** | | | |
|---|---|---|---|---|
| **Ceramide** | **Total effect estimate** | **Total effect p-value** | **ACME estimate** | **ACME p-value** |
| C16 Cer | 2.634e-04 (1.954e-05;5.004e-04) | 0.044 | 7.641e-06 (-1.421e-04;1.796e-04) | 0.908 |
| C18 Cer | 5.527e-04 (-1.182e-04;1.259e-03) | 0.102 | 4.530e-05 (-3.698e-04;4.983e-04) | 0.798 |
| C20 Cer | -2.777e-05 (-7.603e-05;2.105e-05) | 0.314 | 2.285e-05 (-1.929e-06;5.199e-05) | 0.076 |
| C22 Cer | 1.432e-04 (-1.988e-04;4.755e-04) | 0.37 | 2.973e-05 (-1.230e-04;2.119e-04) | 0.788 |
| C24 Cer | 2.703e-02 (-9.487e-03;6.170e-02) | 0.168 | -1.562e-03 (-2.448e-02;1.971e-02) | 0.88 |
| C24:1 Cer | 1.178e-02 (-2.336e-03;2.387e-02) | 0.094 | -9.104e-04 (-8.647e-03;6.183e-03) | 0.85 |
| | **HbA1c** | | | |
| **Ceramide** | **Total effect estimate** | **Total effect p-value** | **ACME estimate** | **ACME p-value** |
| C16 Cer | 2.634e-04 (1.248e-05;4.898e-04) | 0.026 | 4.980e-05 (-8.159e-05;9.854e-05) | 0.298 |
| C18 Cer | 5.527e-04 (-1.136e-04;1.177e-03) | 0.092 | 2.454e-04 (-1.558e-04;3.916e-04) | 0.218 |
| C20 Cer | -2.777e-05 (-7.732e-05;2.150e-05) | 0.272 | 2.416e-06 (-7.150e-06;3.659e-05) | 0.686 |
| C22 Cer | 1.432e-04 (-1.994e-04;4.657e-04) | 0.384 | -5.123e-05 (-1.410e-04;2.217e-05) | 0.14 |
| C24 Cer | 2.703e-02 (-1.441e-02;6.561e-02) | 0.172 | 1.250e-02 (-1.552e-02;2.195e-02) | 0.402 |
| C24:1 Cer | 1.178e-02 (-1.764e-03;2.417e-02) | 0.084 | 4.853e-03 (-4.723e-03;8.500e-03) | 0.354 |
| | **Log10MeanUAER** | | | |
| **Ceramide** | **Total effect estimate** | **Total effect p-value** | **ACME estimate** | **ACME p-value** |
| C16 Cer | 2.883e-04 (4.133e-05;5.397e-04) | 0.02 | -4.061e-05 (-1.156e-04;3.403e-05) | 0.264 |
| C18 Cer | 6.447e-04 (-5.754e-05;1.316e-03) | 0.076 | -8.054e-05 (-3.151e-04;7.562e-05) | 0.332 |
| C20 Cer | -3.459e-05 (-9.307e-05;1.941e-05) | 0.234 | 1.876e-05 (-1.824e-06;4.053e-05) | 0.066 |
| C22 Cer | 1.308e-04 (-2.038e-04;4.675e-04) | 0.428 | 8.857e-06 (-6.075e-05;1.160e-04) | 0.784 |
| C24 Cer | 3.075e-02 (-6.074e-03;6.806e-02) | 0.096 | -3.568e-03 (-1.669e-02;4.336e-03) | 0.346 |
| C24:1 Cer | 1.357e-02 (-2.218e-04;2.626e-02) | 0.056 | -8.651e-04 (-4.897e-03;2.050e-03) | 0.558 |

Mediation effect was assessed by linear regression models with and without adjustment for the tested confounder the effect and significance were estimated by bootstrapping 1000 times using the mediator package in R. ACME:  Average casual mediation effect. UAER: Urinary albumin excretion rate.
